# Individual Differences in Sensitivity to Daily Meteorological Factors Among Collegiate Baseball Players: A Multivariable Repeated-Measures Analysis of Physical Condition and Performance

**DOI:** 10.64898/2026.01.29.26345011

**Authors:** Koji Miyashita

**Affiliations:** Department of Physical Therapy, Chubu University, Aichi, Japan

## Abstract

**Background:** Meteorological factors such as barometric pressure, humidity, and temperature have been linked to weather-related symptoms in the general population, yet little is known about their influence on athletes’ daily well-being and performance. Individual variability in weather sensitivity has been reported in biometeorology research, suggesting that only certain individuals exhibit pronounced physiological responses to environmental fluctuations. However, no studies have examined within-person associations between multiple meteorological factors and daily condition or performance in competitive athletes.

**Methods:** Collegiate baseball players were monitored over 10 randomly selected days during July–August 2025. Subjective condition and performance were assessed daily using a 3-point Likert scale (1 = poor, 2 = normal, 3 = good). Barometric pressure, humidity, and temperature were recorded hourly and summarized for each day using mean values, day-to-day changes, daily ranges, and rapid fluctuation indices. For each player, multivariable linear regression models were constructed to examine within-person associations between the three meteorological variables and daily condition or performance. Model fit (R^2^), regression coefficients (β), and dominant meteorological factors were extracted.

**Results:** Eighty players were included in the condition model and eighty-six in the performance model. High weather sensitivity (R^2^ ≥ 0.60) was observed in 22.5% of players for condition and 14.0% for performance, whereas low sensitivity (R^2^ ≤ 0.20) was found in 26.3% and 16.3%, respectively. Temperature was the dominant explanatory factor in more than 80% of players, although subsets showed dominance of barometric pressure or humidity. Directionality varied across individuals: decreases in barometric pressure were associated with worsening conditions in 62.5% of players but improvement in 37.5%; similar bidirectional patterns were observed for humidity and temperature.

**Conclusion:** Daily meteorological fluctuations explain a meaningful proportion of within-person variation in condition and performance for a subset of collegiate baseball players. The substantial individual variability and diverse directional responses highlight weather sensitivity as a personalized characteristic rather than a uniform effect. These findings suggest that meteorological factors may represent a relevant contextual variable for daily readiness monitoring in susceptible athletes.

## Introduction

Daily fluctuations in meteorological conditions, including barometric pressure, humidity, and temperature, are known to influence a range of human physiological and psychological responses. Within the field of biometeorology, weather-related symptoms such as headache, joint discomfort, fatigue, and mood changes have been widely documented, and increasing attention has been directed toward “weather sensitivity,” in which certain individuals exhibit heightened responses to environmental changes. Previous studies in migraine populations have demonstrated that decreases or rapid fluctuations in barometric pressure can trigger or exacerbate headache symptoms, suggesting that the *change* in atmospheric conditions, rather than the absolute level itself, may play a critical role in symptom generation^1)^. Additionally, epidemiological work on weather-related pain has identified substantial inter-individual variability, with only a subset of individuals responding strongly to relatively small meteorological fluctuations^2)^. These findings collectively highlight the possibility that sensitivity to daily weather changes is not universal and may reflect inherent physiological differences among individuals.

In the context of competitive sports, athletes’ daily condition—reflected in subjective well-being, fatigue, and perceived performance—exhibits considerable day-to-day variability. While training load, recovery status, sleep, and psychological factors have been studied extensively as determinants of readiness and performance, meteorological factors have received comparatively little attention within sports science. Yet environmental ergonomics research has long emphasized that human performance can be modulated by environmental stressors such as heat, humidity, and barometric shifts^3)^. Despite this, few studies have systematically examined whether ordinary meteorological fluctuations influence athletes’ perceived condition during routine training periods. Furthermore, given the documented individual variability in weather sensitivity, it is plausible that only a subathletesletes responds measurably to atmospheric changes, while others remain largely unaffected. Identifying such individual differences may have practical relevance, particularly as subjective measures of well-being are known to be more sensitive than objective metrics in detecting day-to-day variations in athlete readiness^4)^.

To date, no studies have evaluated within-person associations between multiple meteorological factors and both subjective condition and subjective performance in competitive athletes using repeated daily assessments. Therefore, the present study aimed to examine whether daily changes in barometric pressure, humidity, and temperature are associated with self-reported condition and performance among collegiate baseball players. By analyzing each athlete individually using multivariable regression, we sought to characterize the proportion of athletes whose daily condition or performance is meaningfully explained by meteorological factors, to identify which weather elements exert the strongest influence, and to clarify the direction of these associations. This exploratory approach provides foundational evidence regarding individual differences in meteorological sensitivity in competitive athletes and offers insight into how environmental variability may contribute to fluctuations in day-to-day readiness.

## Methods

### Participants

A total of 133 male collegiate baseball players belonging to a university varsity team were recruited. All participants were informed of the study purpose and data confidentiality, and written informed consent was obtained prior to participation. Daily self-report data were collected over a 10-day observation period during July–August 2025. Players who provided at least seven valid daily responses were included in the analysis. Eighty players were eligible for the condition analysis and 86 players for the performance analysis. This study was approved by the Ethics Committee of Chubu University.

### Self-reported condition and performance

Each day during the observation period, players rated their subjective condition and subjective performance using a 3-point Likert scale (“good,” “normal,” “poor”). Responses were converted into numerical values (good = 3, normal = 2, poor = 1). Missing responses were treated as missing data, and days without a report were excluded from statistical analysis.

These subjective indicators were selected because daily well-being measures are known to sensitively reflect short-term fluctuations in athlete readiness and training response^1)^.

### Meteorological data acquisition

Meteorological data were recorded using a temperature–humidity–barometric data logger (TR-73U, T&D Corporation, Nagano, Japan) placed in Kasugai City, Aichi Prefecture, the training location of the team. Barometric pressure (hPa), relative humidity (%), and temperature (°C) were recorded at 1-hour intervals throughout the entire observation period. For each evaluation day, the following daily meteorological indices were calculated for each variable (pressure, humidity, temperature):

1. Daily mean value Mean of the 24-hourly measurements.
2. Day-to-day change (Δ) Difference between the daily mean of the current day and the preceding day.
3. Daily range Difference between the maximum and minimum hourly values within the day.
4. Rapid fluctuation index

Maximum absolute change between consecutive hourly measurements (i.e., max |x_i_ − x_i−1_|). These indices were computationally derived using Python and linked to each player’s self-report on the corresponding day.

### Statistical analysis

#### 1. Individual-level multivariable regression

Because weather responses are known to vary substantially between individuals, analyses were conducted within each athlete rather than at the group level. For each player, two separate multiple linear regression models were constructed:

- Model A (Condition): Condition_i_ = β_0_ + β_1_·Pressure_i_ + β2·Humidity_i_ + β_3_·Temperature_i_ + ε_i_

- Model B (Performance): Performance_i_ = β_0_ + β_1_·Pressure_i_ + β2·Humidity_i_ + β_3_·Temperature_i_ + ε_i_

Players with fewer than 3 valid data pairs or zero variance in the outcome variable were excluded for that specific model. For each model, the following parameters were extracted:

- R^2)^ (variance explained by meteorological factors)
- β coefficients (direction and magnitude of influence)
- Dominant factor (variable with the largest absolute β)

Following conventional interpretations for repeated-measures regression in applied physiology:

- R^2)^ ≥ 0.60 → High weather sensitivity
- R^2)^ ≤ 0.20 → Low or negligible weather sensitivity Intermediate values were also summarized.

#### 2. Classification of dominant meteorological factors

For each athlete, the weather factor with the largest absolute β coefficient was identified as:

- Pressure-dominant
- Humidity-dominant
- Temperature-dominant

The number and proportion of players belonging to each category were summarized.

#### 3. Direction of influence

For each meteorological variable and outcome:

- β > 0 → A *decrease* in the meteorological factor was associated with *worsening* of the outcome
- β < 0 → A *decrease* was associated with *improvement*

The number of players showing positive and negative associations was counted.

#### 4. Software

All data preprocessing and statistical analyses were performed using Python 3.10, with pandas, NumPy, scikit-learn, and stats models packages.

## Results

### Model fit and variance explained by meteorological factors

A total of 80 players were included in the condition model and 86 in the performance model. The distribution of R^2^ values for the individual multivariable regression models is shown in Table 1. For condition, 18 players (22.5%) demonstrated high weather sensitivity (R^2^ ≥ 0.60), indicating that more than 60% of their day-to-day variation in condition was explained by the three meteorological factors. In contrast, 21 players (26.3%) demonstrated low sensitivity (R^2^ ≤ 0.20), suggesting minimal explanatory contribution of weather factors.

**Table 1.** Distribution of R^2^ values for multivariable regression models.

| Outcome | High ($R^2 \geq 0.60$) | Moderate ($0.20 < R^2 < 0.60$) | Low ($R^2 \leq 0.20$) |
| --- | --- | --- | --- |
| Condition | 18 (22.5%) | 41 (51.3%) | 21 (26.3%) |
| Performance | 12 (14.0%) | 60 (69.8%) | 14 (16.3%) |

*Notes: $R^2$ categories define high ( $\geq 0.60$ ), moderate ( $0.20-0.60$ ), and low ( $\leq 0.20$ ) meteorological sensitivity.*

The remaining 41 players (51.3%) fell within the moderate range.

For performance, 12 players (14.0%) exhibited high sensitivity (R^2^ ≥ 0.60), whereas 14 players (16.3%) showed low sensitivity (R^2^ ≤ 0.20). The majority (69.8%) displayed moderate R^2^ values. These findings indicate substantial inter-individual heterogeneity in the degree to which daily meteorological variation explains subjective condition and performance.

This figure shows the distribution of R^2^ values derived from within-person multivariable regression models examining the influence of barometric pressure, humidity, and temperature on daily subjective condition and performance. Athletes demonstrated substantial variability in model fit, with 15–22% showing high weather sensitivity (R^2^ ≥ 0.60) and 16– 26% showing low sensitivity (R^2^ ≤ 0.20). The figure highlights the heterogeneous degree to which meteorological factors explain day-to-day fluctuations across individuals.

### Dominant meteorological factors

Table 2 summarizes the weather factor with the largest absolute regression coefficient (|β|) for each athlete. For both condition and performance, temperature emerged as the dominant factor for more than 80% of players (condition: 81.3%; performance: 81.4%).

**Table 2.** Dominant meteorological factor (largest |β|)

| Outcome | Pressure | Humidity | Temperature |
| --- | --- | --- | --- |
| Condition | 5 (6.3%) | 10 (12.5%) | 65 (81.3%) |
| Performance | 7 (8.1%) | 9 (10.5%) | 70 (81.4%) |

*Notes: Dominant factor is defined as the meteorological variable with the largest absolute regression coefficient ( $|\beta|$ ) within each athlete.*

However, a non-negligible subset of athletes showed dominance of humidity (condition: 12.5%; performance: 10.5%) or barometric pressure (6.3% and 8.1%, respectively). This pattern indicates that the type of meteorological factor influencing daily status varied across individuals and was not uniform within the team.

This heatmap displays the standardized β coefficients for barometric pressure, humidity, and temperature for each athlete. Warmer colors indicate stronger positive associations, whereas cooler colors represent negative associations. The pattern reveals diverse meteorological response profiles, including temperature-dominant, humidity-dominant, and pressure-dominant individuals. Bidirectional associations (positive and negative β) are evident across all variables, indicating individualized sensitivity rather than uniform group-level effects.

### Visualization of regression coefficients

The distribution of standardized β coefficients for temperature and barometric pressure in the condition model is illustrated in Figure 1. Athletes were dispersed across all four quadrants, indicating that both positive and negative response directions existed for each meteorological factor. This pattern visually highlights the substantial inter-individual heterogeneity in meteorological sensitivity and aligns with the numerical findings presented in Tables 2 and 3.

**Table 3.** Direction of associations based on β sign.

| Variable | Decrease → Worse ($\beta > 0$) | Decrease → Better ($\beta < 0$) |
| --- | --- | --- |
| Condition – Pressure | 50 (62.5%) | 30 (37.5%) |
| Condition – Humidity | 40 (50.0%) | 40 (50.0%) |
| Condition – Temperature | 30 (37.5%) | 50 (62.5%) |
| Performance – Pressure | 48 | 38 |
| Performance – Humidity | 36 | 50 |
| Performance – Temperature | 41 | 45 |

*Notes: Positive $\beta$ indicates worsening with decreases in the meteorological factor; negative $\beta$ indicates improvement with decreases.*

**Figure 1.**
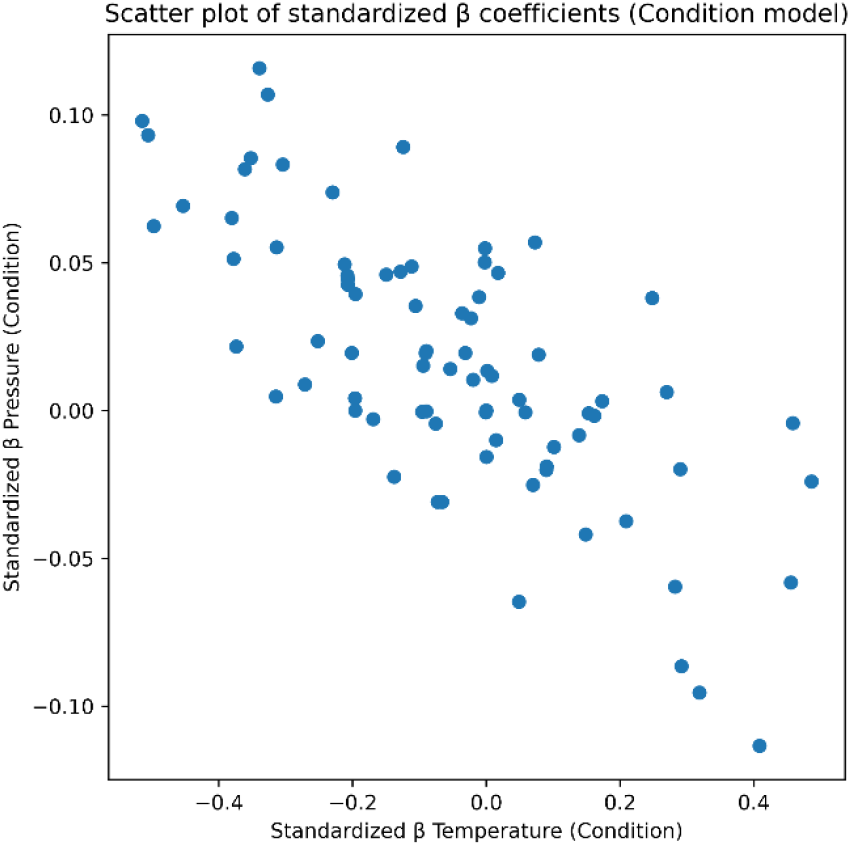
Scatter plot of standardized β coefficients for temperature and barometric pressure in the condition model. Each point represents one athlete. The distribution across all four quadrants indicates substantial inter-individual heterogeneity in both the direction and magnitude of meteorological sensitivity.

An analogous pattern was observed in the performance model (Figure 2), where standardized β coefficients for temperature and barometric pressure were similarly scattered across all quadrants, indicating heterogeneous and bidirectional meteorological influences on perceived performance at the individual level.

**Figure 2.**
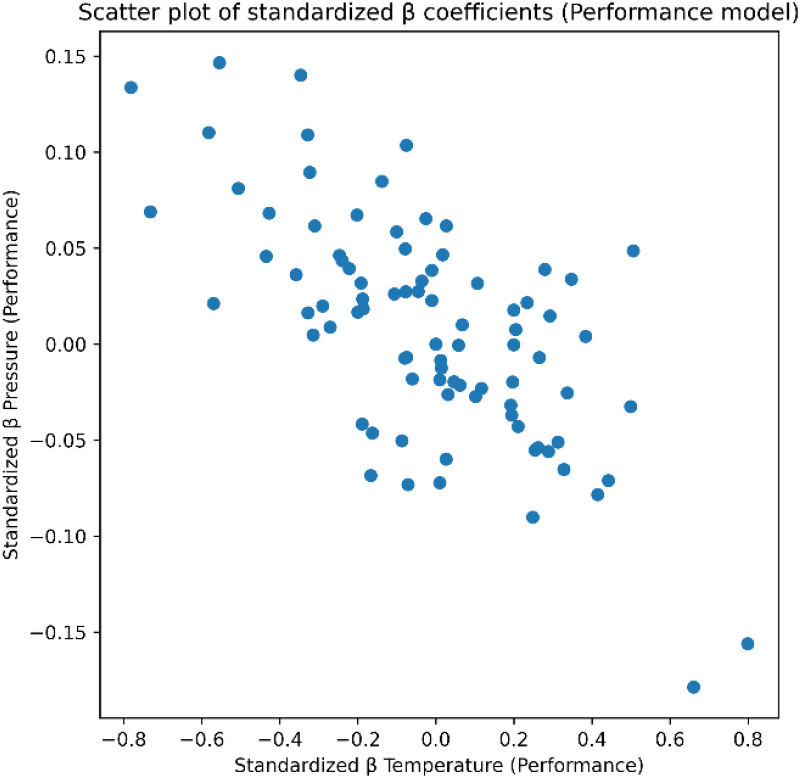
Scatter plot of standardized β coefficients for temperature and barometric pressure in the performance model. Each dot represents one athlete. The wide dispersion across quadrants demonstrates substantial inter-individual variation in meteorological sensitivity related to performance.

### Direction of associations with meteorological factors

Table 3 presents the number of athletes showing positive or negative β coefficients for each meteorological variable. For condition, decreases in barometric pressure were associated with worsening (β > 0) in 50 players (62.5%), whereas 30 players (37.5%) showed the opposite pattern. Humidity exhibited a balanced distribution (β > 0 in 50.0%; β < 0 in 50.0%). For temperature, 30 players (37.5%) showed worsening with decreasing temperature, while the majority (62.5%) showed improvement.

This scatter plot visualizes clusters derived from multivariable regression parameters, representing distinct meteorological sensitivity profiles among athletes. Clusters reflect differences in dominance patterns, β directionality, and overall model fit. The figure illustrates that subsets of athletes share similar weather-response characteristics, while others form unique profiles, further supporting considerable inter-individual variability.

For performance, similar bidirectional patterns were observed. Barometric pressure decreases were associated with worsening performance in 48 players and improvement in 38.Humidity (36 vs. 50) and temperature (41 vs. 45) also showed mixed effects. Together, these results highlight bimodal response patterns, suggesting that meteorological fluctuations may improve daily condition in some athletes while worsening it in others.

### Overall interpretation

Across the sample, weather sensitivity varied widely, with about 15–22% of athletes showing strong meteorological influence and 16–26% showing minimal influence.

Temperature was the most common dominant factor, but meaningful subsets of athletes exhibited barometric-pressure or humidity dominance. In addition, the direction of influence was not uniform; both positive and negative β values were observed across all factors, indicating substantial individual variability in meteorological response patterns.

## Discussion

The present study investigated individual-level associations between daily meteorological fluctuations and subjective condition and performance among collegiate baseball players. Using repeated within-person assessments and multivariable regression models, we demonstrated that a considerable subset of athletes showed moderate to strong relationships between weather factors and day-to-day variations in well-being. These findings provide novel evidence that ordinary, subclinical meteorological changes—occurring within the range of normal weather variability—may represent a meaningful external factor influencing daily readiness in competitive athletes.

### Individual variability in weather sensitivity

One of the most notable findings was the substantial inter-individual variability in weather sensitivity. Approximately 15–22% of athletes exhibited high sensitivity (R^2^ ≥ 0.60), meaning that meteorological factors accounted for more than half of their daily fluctuations in condition or performance. Conversely, 16–26% showed minimal sensitivity (R^2^ ≤ 0.20). This bimodal pattern aligns with prior epidemiological research demonstrating that only specific subgroups exhibit pronounced responses to barometric pressure or other weather variables^1)^,^2)^. Our results extend this concept to competitive athletes and suggest that meteorological sensitivity is not a universal characteristic but rather an individual trait.

### Differential contributions of meteorological factors

Temperature emerged as the dominant explanatory factor for more than 80% of players, whereas humidity or barometric pressure were the dominant variables for a minority. This distribution suggests that thermal stress and its day-to-day fluctuations may exert broader physiological effects across athletes, consistent with the literature on environmental ergonomics^3)^. However, the presence of pressure-dominant or humidity-dominant responders indicates important heterogeneity. Prior research on barometric pressure–related symptoms has shown that sudden or substantial pressure changes can modulate sensory and autonomic activity^1)^,^2)^.

Furthermore, experimental neurophysiological studies have demonstrated that rapid decreases in barometric pressure increase neuronal activity within trigeminal nociceptive pathways, providing a mechanistic explanation for weather-associated symptoms^5)^. Such mechanisms— mediated through autonomic reactivity or altered sensory processing—may also contribute to the individual differences in meteorological sensitivity observed in athletes.

### Bidirectional effects and complex response patterns

An additional novel finding is that the direction of meteorological influence was not uniform across players. For every weather variable, both positive and negative β coefficients were observed. For example, decreases in barometric pressure were associated with worsening conditions in some players while improving conditions in others. These bidirectional patterns suggest that meteorological factors may interact with individual physiological baselines, lifestyle routines, recovery status, or genetic predispositions. Such heterogeneity resonates with the broader sports science literature demonstrating substantial individual differences in responses to standardized training loads^4)^.

### Graphical confirmation of heterogeneous response patterns

As depicted in Figures 1 and 2, the β coefficients for temperature and barometric pressure in the condition and performance models were widely distributed across all directional quadrants. This dispersion reinforces the presence of multiple meteorological response phenotypes—temperature-dominant, pressure-dominant, humidity-influenced, and mixed-response athletes—and supports the interpretation that meteorological sensitivity is a highly individualized characteristic rather than a uniform population-level effect.

### Implications for athlete monitoring

Subjective measures of well-being are among the most sensitive indicators for detecting daily changes in athlete readiness, often outperforming objective markers such as heart-rate variability or neuromuscular tests.^6)^,^7^ Prior research has shown that self-reported wellness scales can respond more rapidly to training-induced fluctuations than objective physiological measures, reinforcing the value of subjective monitoring in applied sport settings.^6)^,^7)^

The present study supports the utility of these subjective assessments by showing that they captured subtle fluctuations associated with environmental conditions. For athletes with high meteorological sensitivity, incorporating weather-aware monitoring may enhance interpretation of daily fatigue, readiness, and perceived performance trends. Conversely, for athletes with low sensitivity, such factors may be less relevant compared with internal load or psychological indicators.

### Methodological contribution

To our knowledge, this is the first study to analyze multivariable meteorological influences using repeated within-person models in athletes, simultaneously incorporating barometric pressure, humidity, and temperature. This analytic approach avoids group-averaging artifacts and highlights individual response profiles, offering a more precise understanding of how environmental factors relate to subjective condition in real-world athletic settings.

### Limitations

This study has several limitations. First, the observation period was limited to 10 days, which constrains the stability and reliability of estimated regression coefficients. Longer-term monitoring across different seasons is required to determine whether meteorological sensitivity is consistent over time.

Second, although multivariable regression was used to model within-person effects, the study design does not permit causal inference. Daily condition and performance may be influenced by numerous factors—including training load, sleep quality, psychological stress, hydration status, and recovery practices—that were not measured in this study.

Third, meteorological variables were recorded at a single location near the training environment, and individual microclimates or indoor/outdoor exposure differences were not assessed.

Fourth, only subjective outcomes were analyzed. Although subjective measures are highly sensitive to day-to-day variation, complementary objective indicators (e.g., physical performance tests, heart-rate variability, neuromuscular metrics) would strengthen interpretation.

Finally, the sample consisted of a single collegiate baseball team, limiting generalizability to other sports, competitive levels, or age groups. Future studies incorporating larger and more diverse athletic populations, additional environmental factors, and longer monitoring periods are warranted to more comprehensively characterize meteorological influences on athlete readiness.

## Conclusion

This study demonstrated that daily fluctuations in meteorological factors—specifically barometric pressure, humidity, and temperature—exhibit measurable associations with subjective condition and performance in a subset of collegiate baseball players. Using repeated within-person multivariable regression, we identified substantial individual variability in meteorological sensitivity, with approximately 15–22% of athletes showing high sensitivity (R^2^ ≥ 0.60) and 16–26% showing minimal sensitivity (R^2^ ≤ 0.20). Temperature was the most common dominant factor, while barometric pressure and humidity also influenced certain individuals. The direction of associations varied across athletes, indicating bidirectional response patterns rather than uniform effects.

These findings suggest that ordinary meteorological variability may represent a meaningful contextual factor influencing day-to-day readiness in some athletes, and that simple subjective self-report measures can capture these relationships. Identifying athletes who are sensitive to specific weather elements may contribute to more individualized interpretation of daily wellness data and improved understanding of fluctuations in perceived performance.

## Data Availability

Not applicable

## References

1. Okuma H, Okuma Y, Kitagawa Y. Examination of fluctuations in atmospheric pressure related to migraine. Internal Medicine. 2015;54(4):435–439.

2. Sato J, et al. Epidemiology and clinical characteristics of weather-related pain. Pain Research. 2021;36(2):1–8.

3. Parsons KC. Environmental ergonomics: A review of principles, methods and models. Applied Ergonomics. 2000;31(6):581–594.

4. Mann TN, Lamberts RP, Lambert MI. High responders and low responders: Factors associated with individual variation in response to standardized training. Sports Medicine. 2014;44(8):1113–1124.

5. Messlinger K, Funakubo M, Sato J, Mizumura K. Increases in neuronal activity in the rat spinal trigeminal nucleus following changes in barometric pressure: Relevance for weather-associated headaches? Headache. 2010;50(9):1449–1463.

6. Saw AE, Main LC, Gastin PB. Monitoring the athlete training response: Subjective self-reported measures trump commonly used objective measures. Br J Sports Med. 2016;50(5):281–289.

7. Hooper SL, Mackinnon LT. Monitoring overtraining in athletes: Recommendations. Sports Medicine. 1995;20(5):321–327.

